# In-flight Transmission Cluster of COVID-19: A Retrospective Case Series

**DOI:** 10.1101/2020.03.28.20040097

**Authors:** Naibin Yang, Yuefei Shen, Chunwei Shi, Ada Hoi Yan Ma, Xie Zhang, Xiaomin Jian, Liping Wang, Jiejun Shi, Chunyang Wu, Guoxiang Li, Yuan Fu, Keyin Wang, Mingqin Lu, Guoqing Qian

**Affiliations:** Department of Infection and Liver Diseases, Ningbo First Hospital, Ningbo 315010, China; Department of Infectious Diseases, Xiaoshan First People’s Hospital, Xiaoshan 311200, China; Nottingham University Business School, University of Nottingham Ningbo China, Ningbo315010, China; Department of Pulmonary and Critical Care Medicine, Yueqing people’s Hospital, Yueqing 325600, China; Department of Radiology, Beijing Fuxing Hospital Affiliated to Capital University of Medical Science, Beijing 100038, China; Department of Radiology, Ningbo First Hospital, Ningbo 315010, China; Department of Infection and Liver Diseases, the First Hospital of Jiaxing City, Jiaxing 314000, China; Department of Infection and Liver Diseases, the first affiliated hospital of Wenzhou Medical University, Wenzhou 325000, China; Department of General Internal Medicine, Ningbo First Hospital, Ningbo 315010, China

**Keywords:** COVID-19, SARS-CoV-2, aircraft, passenger, flight

## Abstract

**Objectives:** No data were available about in-flight transmission of SARS-CoV-2. Here, we report an in-flight transmission cluster of COVID-19 and describe the clinical characteristics of these patients.

**Methods:** After a flight, laboratory-confirmed COVID-19 was reported in 12 patients. Ten patients were admitted to the designated hospital. Data were collected from 25^th^ January to 28^th^ February 2020. Clinical information was retrospectively collected.

**Results:** All patients are passengers without flight attendants. The median age was 33 years, and 70% were females. None was admitted to intensive care unit, and no patients succumbed through 28^th^ February. The median incubation period was 3.0 days and from illness onset to hospital admission was 2 days. The most common symptom was fever. Two patients were asymptomatic and negative for chest CT scan throughout the disease courses. On admission, initial RT-PCR were positive in 9 patients, however initial chest CT were positive in only half patients. The median lung “total severity score” of chest CT was 6. Notably, “Crazy-Paving” pattern, pleural effusion, and ground-glass nodules were also seen.

**Conclusion:** It is potential for COVID-19 transmission by airplane, but the symptoms are mild. Passengers and attendants must be protected during the flight.

## Introduction

Since the initial pneumonia cases of unknown cause in early December 2019 were reported [1] and subsequently confirmed to be the novel coronavirus disease (COVID-19) caused by a novel coronavirus named severe acute respiratory syndrome coronavirus 2 (SARS-CoV-2) [2], this emerging infectious disease spread all over China rapidly and many other countries and regions, especially in Europe, Republic of Korea, and Iran. WHO made the assessment that COVID-19 spread should be characterized as a pandemic on 12^th^ March, 2020.

The initial outbreak of COVID-19 in Wuhan and other cities in China was triggered because of respiratory and contact precautions were not in place, and people frequently participate in gatherings before the Chinese lunar new year holiday. It was reported that even brief encounters with COVID-19 patients may lead to infection. Even an asymptomatic person can lead a family clustering of SARS-CoV-2 infection[3]. COVID-19 can be transmitted from human to human through respiratory droplets, contact with infected persons and possibly via fecal-oral transmission [4]. COVID-19 has a feature of family or social cluster, and transmission while using public transport.

COVID-19 is characterized by the acute onset of fever with or without cough, fatigue, shortness of breath, and sometimes diarrhea. The incubation period had an average of 3 days, longer up to 24 days [4]. Most cases were mild, but serious cases might suffer severe pneumonia, acute respiratory distress syndrome (ARDS) and multiple organ failures, eventually leading to death[5]. The mortality rate of confirmed cases up to 12^th^ Mar 2020 in China were 3.93% (3181/81007), higher than that (3.39%, 1886/55471) in cases reported by other countries outside[6].

After the implementation of control measures applied to prevent further spread in China, the spread was eventually contained. These measures include banning gatherings, isolation of suspected persons, the great efforts for effective diagnosis and treatment, daily public educational campaigns on precautionary measures that each person can take to avoid getting exposed to SARS-CoV-2[7]. As of 12^th^ March, within China, for many days there had been fewer than 10 newly confirmed cases per day. However, outbreak of COVID-19 outside of China is gathering pace with rapidly increasing numbers of new confirmed cases. As of 12^th^ March more than 50000 cases have been reported[6]. Further rapid increases are expected to continue for some time.

The COVID-19 spreads rapidly around the world, largely because of persons infected with the SARS-CoV-2 traveled on aircraft to other countries, since SARS-Cov-2 has strong affinity to human respiratory receptors[8]. No data were available about the risk of SARS-CoV-2 transmission on aircraft and clinical characteristics and outcomes of these COVID-19 patients. Here, we conducted a retrospective research to describe the clinical characteristics in an attempt to recognize the features of COVID-19 infected on aircraft.

## Methods

### Background

Since the early outbreak of COVID-19 in China, strict precautionary measures are implemented. Especially, Zhejiang provincial government initially launched a Category A response – a response to a major public health emergency – in the morning of 23^th^ January 2020. In one day, a commercial aircraft carrying 325 passengers and crew members flew to Hangzhou International Airport in Zhejiang. Most passengers were Chinese tourists and were returning home. When they were boarding, the work group in Singapore airport implemented strict preflight screening to find out whether they had fever or respiratory symptoms. None of them were symptomatic. Most passengers had taken no precautionary measures against possible exposure to SARS-CoV-2, while all the flight attendants well wore masks. During the flight, a man passenger (patient 1) who traveled from Wuhan had a fever and no respiratory symptoms. Also, he did not wear a mask. After the 5-hour flight, the plane arrived in Hangzhou at 10 pm. Considering there were some Wuhan citizens on aircraft, staffs from the Hangzhou Center for Disease Control and Prevention (CDC) had been on standby at the airport and once the aircraft arrived they boarded to screen for possible fever and respiratory symptoms amongst the passengers and crew members. They had found that patient 1 still had a fever and his temperature was 38.1□. Then, his throat swab specimens were collected and sent to CDC. Five hours later, he was confirmed positive for SARS-CoV-2 and was admitted into the designated hospital. The crew members and rest of the passengers were placed under isolation and routine medical checks at hotels near airport for 14 days to observe whether illness onset will happen. As shown in figure 1, a further nine passengers were successively tested positive for SARS-CoV-2 and were also admitted. Another two passengers were also tested positively but we failed to interview them. No further patients were found in the two weeks of isolation among the remaining 313 passengers and crew members. Then they were released from quarantine.

**Figure 1:**
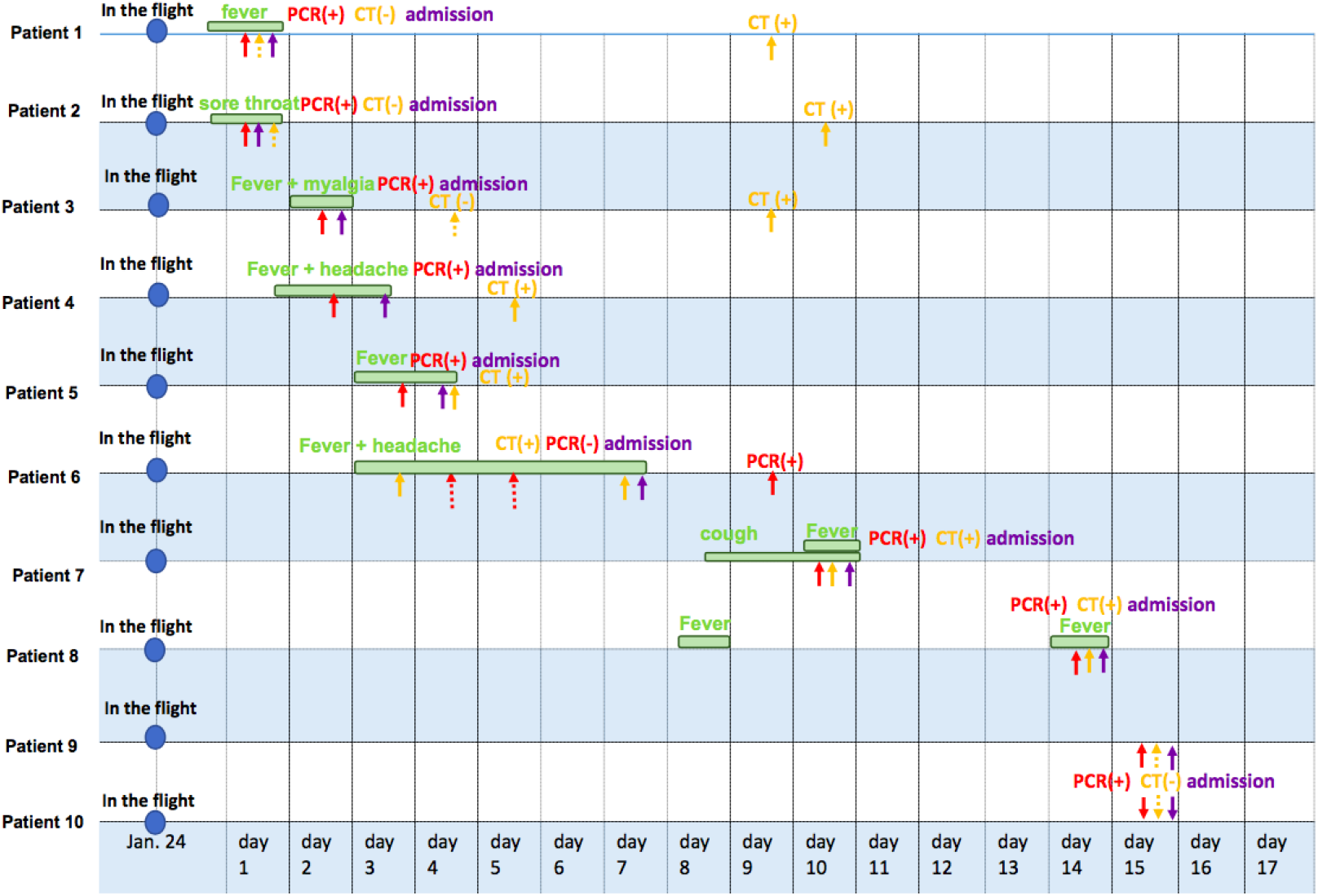
Timeline of onset of illness, diagnosis, and admission in ten patients with COVID-19 by transmission on aircraft (The periods from symptoms onset to admission were marked by green column; timepoints were markedly with RT-PCR positive by solid red arrow and RT-PCR negative by virtual red arrow; timepoints were markedly with Chest CT positive by solid yellow arrow and chest CT negative by virtual yellow arrow; dates of admission were marked by solid purple arrow)

### DATA sources

We conducted a retrospective study focused on the clinical characteristics of ten patients with COVID-19 successively admitted to Xiaoshan First People’s Hospital after having been on a flight. Patients were diagnosed based on National Health Commission of the People’s Republic of China guidelines and the WHO interim guidance[9, 10]. For the ten patients included in our study, there was no recognized exposure history in 14 days before this fight. The incubation period was defined as the time from the flight arrival day to the onset of illness. Hospitalized days were defined as the period from admission to discharge day. The study has been reviewed and approved by the Medical Ethical Committees of Xiaoshan First people’s Hospital (Approval NO: 2020-02). Written informed consent were waived because of the urgent need to collect clinical data on this emerging disease and entirely anonymized data therefore the individual identity cannot be identified.

Epidemiology data were collected by interviewing each patient including exposure history, dates of illness onset, hospital admissions and discharged. Clinical information, including symptoms, chest CT images and therapy during hospital stay, laboratory results on admission, were retrospectively collected from their medical records. Two experienced radiologists (Y. Fu; X. Jian) reviewed all chest CT images and positive/negative findings and radiological features were decided by consensus. Both epidemiology data and clinical information were sent to two experienced physicians (G. Qian; N. Yang). If information were unclear or missing, we clarified the details with the attendant doctors who were treating these patients.

### Laboratory confirmation and treatment

Throat swab specimens collected from all patients were sent to CDC and tested for SARS-CoV-2 RNA by RT-PCR using the established sets of primers[11]. Outcomes were ready in three hours. Laboratory confirmation was achieved when the result of RT-PCR was positive.

Laboratory tests conducted on the day of admission included blood cell count, C-reactive protein, procalcitonin, serum chemistry, D-dimer, fibrinogen in coagulation test, liver and renal function, electrolytes and lactate dehydrogenase. Each of five lung lobes were assessed for degree of involvement and classified as none (0%) with a score of 0, minimal (1-25%) with a score of 1, mild (26-50%) with a score of 2, moderate (51-75%) with a score of 3, or severe (76-100%) with a score of 4. An overall lung “total severity score” was reached by summing the five lobes scores (range of possible scores, 0-20)[12].

All Patients received antiviral treatment with Arbidol (200mg twice daily, in ten patients), either in combination with Lopinavir/Ritonavir (400mg/100mg twice daily, in seven patients) or Darunavir/Cobicistat (800mg/150mg once daily in three patients). Patients were treated with corticosteroid (40-80mg) when their resting respiratory rate were more than 30 per minutes or when multiple pulmonary lobes showed more than 50% progression of disease on chest CT imaging in two days. Oxygen supports via nasal cannula were given when oxygen saturations were below 93% without oxygen support. Patients were discharged from hospital when the results of two RT-PCR tests taken 24 hours apart were negative for SARS-CoV-2 and symptoms improved obviously according to China guidelines^10^. MuLBSTA scores were assessed and recorded by attending physicians based on six indexes [13].

### Statistical analysis

We summarized continuous variables as medians (interquartile ranges, IQR). For categorical variables, we present the counts and percentages in each category. All analyses were conducted with IBM SPSS statistics software, version 26.0.

## Results

### Demographic and clinical characteristics

By 28^th^ February, clinical data had been collected on all ten patients with laboratory-confirmed COVID-19. No flight attendants have been infected. In 14 days prior to the flight, no one including patient 1 had been exposed to confirmed or suspected COVID-19 patients.

As shown in Table 1, the median incubation period was 3.0 (IQR, 2-7; ranged from 1 to 14 or longer) days and from illness onset to hospital admission was 2 (IQR, 1-4; ranged from 1 to 6) days. The median age was 33 (IQR, 26 to 42; ranged from 20 to 52), and 70% were females and none was pregnant. None of them was health worker.

**Table 1:**
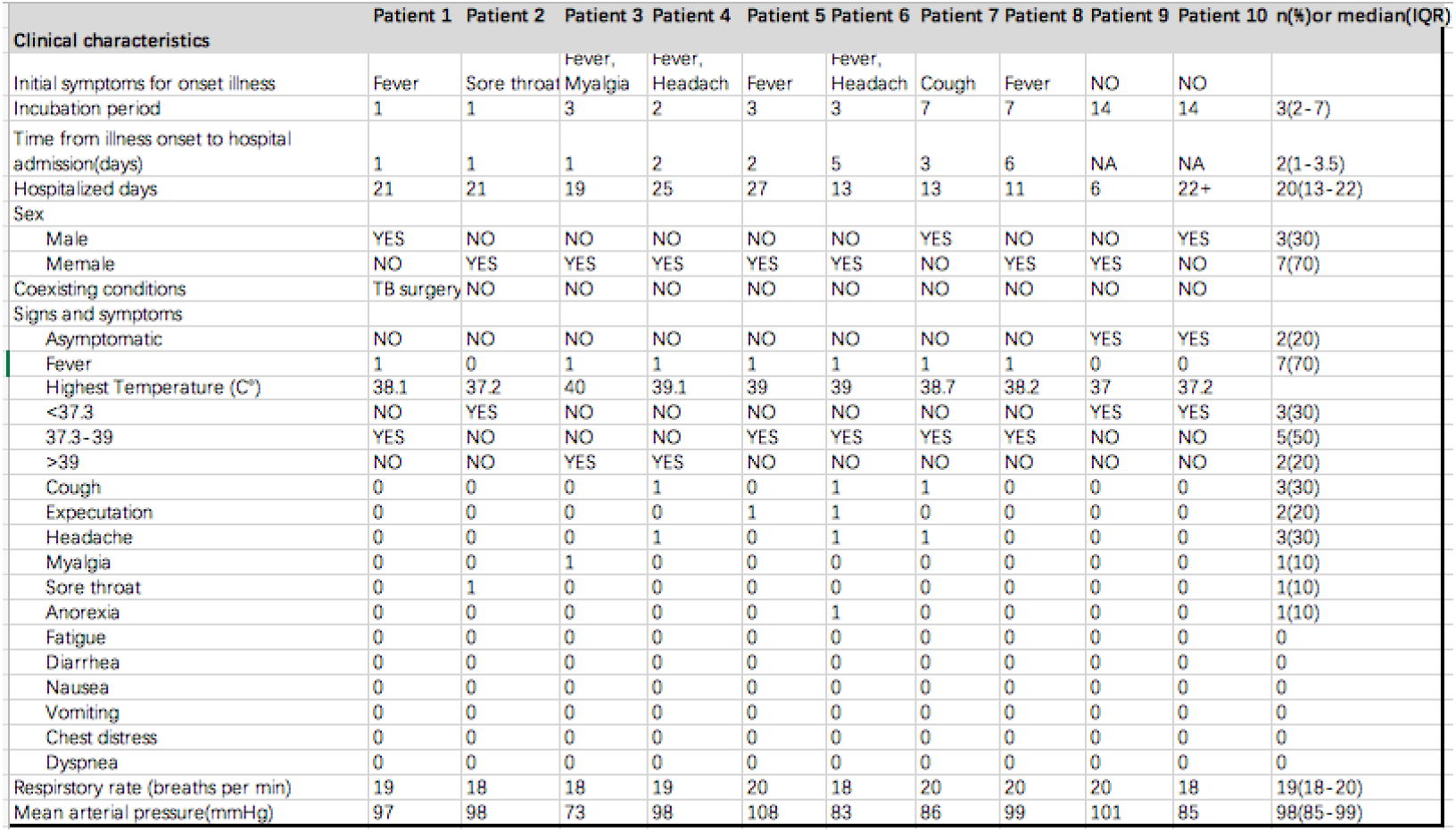
Clinical characteristics of patients with COVID-19 by transmission on aircraft

Of the ten patients, two patients (patient 9 and 10) were asymptomatic and negative for chest CT scan throughout the disease course. In the other eight patients, the most common symptoms were fever in 7 (70%) patients. Among them, 5 (50%) patients had temperature between 37.3-39□, while 2 (20%) patients were above 39□. Other symptoms included cough (3, 30%), headache (3, 30%), expectoration (2, 20%), whereas sore throat, anorexia and myalgia was seen in 1 (10%) patient.

### Laboratory findings on admission

Table 2 shows the laboratory findings on admission. Elevated C-reactive protein (>4mg/L) was seen in 7 (70%) patients, while leucopenia (white blood cell count <3.5⨯10^9^/L) and lymphopenia (lymphocyte count <1.1×10^9^/L) was observed in 1 (10%) patient, respectively. Decreased platelet levels (<125×10^9^/L) were detected in 2 (20%) patients. All the procalcitonin levels were within normal range at less than 0.5μg/L. No patient had an increased level of aspartate aminotransferase. Decreased level of potassium with 2.5mmol/L and increased level of D-dimer with 1.78mg/L were documented in 1 (10%) patient. Two (20%) patients had slight increased levels of fibrinogen (normal range, 2-4g/L).

**Table 2:** Laboratory characteristics of patients with COVID-19 by transmission on aircraft

|  | Patient 1 | Patient 2 | Patient 3 | Patient 4 | Patient 5 | Patient 6 | Patient 7 | Patient 8 | Patient 9 | Patient 10 | n(%)or median(IQR) |
| --- | --- | --- | --- | --- | --- | --- | --- | --- | --- | --- | --- |
| **Laboratory characteristics** |  |  |  |  |  |  |  |  |  |  |  |
| Leukocyte count (x10<sup>9</sup>/L cells/L) | 5.78 | 2.92 | 4.48 | 4.44 | 5.67 | 4.32 | 6.02 | 3.83 | 10.11 | 6.53 | 5.08(4.35-5.96) |
| Normal leukocyte count (3.5-9.5x10<sup>9</sup> cells/L) | YES | NO | YES | YES | YES | YES | YES | YES | NO | YES | 8(80) |
| Low leukocyte count (<3.5x10<sup>9</sup> cells/L) | NO | YES | NO | NO | NO | NO | NO | NO | NO | NO | 1(10) |
| Lymphocyte count (x10<sup>9</sup> cells/L) | 1.16 | 1.04 | 0.67 | 1.1 | 1.57 | 1.35 | 1.93 | 1.26 | 4.38 | 2.44 | 1.31(1.12-1.84) |
| Lympho peria(<1.1x10<sup>9</sup> cells/L) | NO | NO | YES | NO | NO | NO | NO | NO | NO | NO | 1(10) |
| Neutrophil count(x10<sup>9</sup> cells/L) | 3.79 | 1.56 | 2.87 | 2.92 | 3.4 | 2.36 | 3.82 | 2.04 | 4.97 | 3.59 | 3.16(2.49-3.74) |
| C-reactive protein (mg/L) | 4.2 | 5.98 | 2.18 | 5.27 | 1.9 | 10.49 | 47 | 5.66 | 4.2 | 2 | 4.74(2.67-5.9) |
| Elevated C-reactive protein(>4mg/L) | YES | YES | NO | YES | NO | YES | YES | YES | YES | NO | 7(70) |
| Procalcitonin(ug/L, normal: <0.5ug/L) | NA | 0.04 | 0.05 | 0.03 | 0.03 | NA | 0.12 | NA | 0.02 | 0.03 | 0.03(0.03-0.05) |
| Haemoglobin(g/L) | 149 | 125 | 127 | 117 | 143 | 142 | 163 | 124 | 143 | 150 | 143(126-148) |
| Platelet count (x10<sup>9</sup> cells/L) | 196 | 191 | 196 | 279 | 213 | 98 | 114 | 155 | 162 | 233 | 194(157-209) |
| Decreased Platelet(<125x10<sup>9</sup> cells/L) | NO | NO | NO | NO | NO | YES | YES | NO | NO | NO | 2(20) |
| Elevated ALT(>50U/L) or AST (>40U/L) | NO | NO | NO | NO | NO | NO | NO | NO | NO | NO | 0 |
| ALT(U/L) | 17 | 5 | 8 | 20 | 12 | 25 | 23 | 6 | 11 | 12 | 12(9-19) |
| AST(U/L) | 19 | 14 | 10 | 17 | 16 | 25 | 27 | 18 | 15 | 16 | 17(15-19) |
| Creatine (umol/L) | 94 | 57 | 58 | 61 | 65 | 74 | 84 | 63 | 53 | 84 | 64(59-81) |
| Lactate dehydrogenase (U/L) | 144 | 146 | 153 | 182 | 124 | 247 | 264 | 126 | 111 | 135 | 145(128-175) |
| Elevated lactate dehydrogenase: >225 (U/L) | NO | NO | NO | NO | NO | YES | YES | NO | NO | NO | 2(20) |
| Potassium(mmol/L, normal: 3.5-5.3mmol/L) | 4.23 | 4.18 | 2.5 | 3.88 | 4.15 | 4.23 | 3.7 | 4 | 4.2 | 4.2 | 4.17(3.91-4.2) |
| Sodium(mmol/L, normal:137-147mmol/L) | 142 | 141 | 139 | 137 | 140 | 139 | 139 | 143 | 141 | 143 | 141(139-142) |
| Chloridion(mmol/L, normal:99-110mmol/L) | 103.7 | 104.8 | 105.7 | 101.9 | 103.6 | 102.5 | 100.4 | 107.4 | 103.4 | 106.4 | 103.7(102.7-105.5) |
| D-dimer(mg/L) | 0.04 | NA | NA | 0.2 | 0.12 | 0.56 | 0.34 | 0.09 | 1.78 | 0.46 | 0.27(0.11-0.49) |
| Fibrinogen | 2.67 | NA | 2.54 | 3.16 | 4.03 | 3.16 | 4.73 | 3.43 | 1.57 | 2 | 3.16(2.54-3.43) |

### Chest CT findings and the relation with RT-PCR

As shown in table 3, two of the ten patients had no pneumonia throughout the disease course. Typical CT findings of pulmonary parenchymal ground-glass opacities, especially with peripheral lung distribution, were seen in the other 8 patients. Among of them, bilateral lung involvements were seen in 7 patients while unilateral left lung involvement was seen in 1 patient. The most frequently involved lobe was left lower lobe in all the above 8 (80%) patients and right lower lobe in 7 (70%) patients. Of the 7 patients with CT images of bilateral pulmonary parenchymal ground-glass opacities, consolidations were also seen in 4 patients. Notably, “Crazy-Paving” pattern, pleural effusion (Figure 2), and ground-glass nodules were also seen in some patients.

**Table 3:** Imaging characteristics of patients with COVID-19 by transmission on aircraft

|  | Patient 1 | Patient 2 | Patient 3 | Patient 4 | Patient 5 | Patient 6 | Patient 7 | Patient 8 | Patient 9 | Patient 10 | n(% or median(IQR)) |
| --- | --- | --- | --- | --- | --- | --- | --- | --- | --- | --- | --- |
| **Chest CT and RT-PCR** |  |  |  |  |  |  |  |  |  |  |  |
| Quantitative RT-PCR positive | YES | YES | YES | YES | YES | YES | YES | YES | YES | YES | 10(100) |
| Initial RT-PCR positive | YES | YES | YES | YES | YES | NO | YES | YES | YES | YES | 9(90) |
| Chest CT imaging: positive | YES | YES | YES | YES | YES | YES | YES | YES | NO | NO | 8(80) |
| Initial chest CT imaging: positive | NO | NO | NO | YES | YES | YES | YES | YES | NO | NO | 5(50) |
| Positive: RT-PCR>chest CT | YES | YES | YES | NO | NO | NO | NO | NO | YES | YES | 5(50) |
| Positive: RT-PCR<chest CT | NO | NO | NO | NO | NO | YES | NO | NO | NO | NO | 1(10) |
| Bilateral Lung Involvement | YES | YES | YES | YES | YES | YES | YES | NO | NO | NO | 7(70) |
| Unilateral lung involvement | NO | NO | NO | NO | NO | NO | NO | YES | NO | NO | 1(20) |
| No lung lobe Distribution | NO | NO | NO | NO | NO | NO | NO | NO | YES | YES | 2(20) |
| Ground-glass opacities | YES | YES | YES | YES | YES | YES | YES | YES | NO | NO | 8(80) |
| Consolidation | NO | NO | YES | YES | NO | YES | YES | NO | NO | NO | 4(40) |
| Nodules | NO | NO | NO | YES | NO | NO | NO | YES | NO | NO | 2(20) |
| "Crazy-Paving" Pattern | NO | NO | NO | NO | YES | NO | NO | NO | NO | NO | 1(10) |
| Pleural Effusion | NO | NO | NO | NO | YES | NO | YES | NO | NO | NO | 1(10) |
| Pulmonary fibrosis | NO | NO | NO | YES | YES | NO | NO | NO | NO | NO | 2(20) |
| Lobes numbers of with Lung Opacities involvement | 4 | 2 | 5 | 5 | 4 | 5 | 5 | 1 | 0 | 0 | 4(1-5) |
| Lung "total severity score" in CT | 5 | 2 | 9 | 13 | 10 | 8 | 7 | 1 | 0 | 0 | 6(1-9) |
| Involvement of Right Upper Lobe | NO | NO | YES | YES | YES | YES | YES | NO | NO | NO | 5(50) |
| Involvement of Right Middle Lobe | YES | NO | YES | YES | NO | YES | YES | NO | NO | NO | 5(50) |
| Involvement of Right Lower Lobe | YES | YES | YES | YES | YES | YES | YES | NO | NO | NO | 7(70) |
| Involvement of Left Upper Lobe | YES | NO | YES | YES | YES | YES | YES | NO | NO | NO | 6(60) |
| Involvement of Left Lower Lobe | YES | YES | YES | YES | YES | YES | YES | YES | NO | NO | 8(80) |
| Peripheral Distribution | YES | YES | YES | YES | YES | YES | YES | YES | NO | NO | 8(80) |

**Figure 2:**
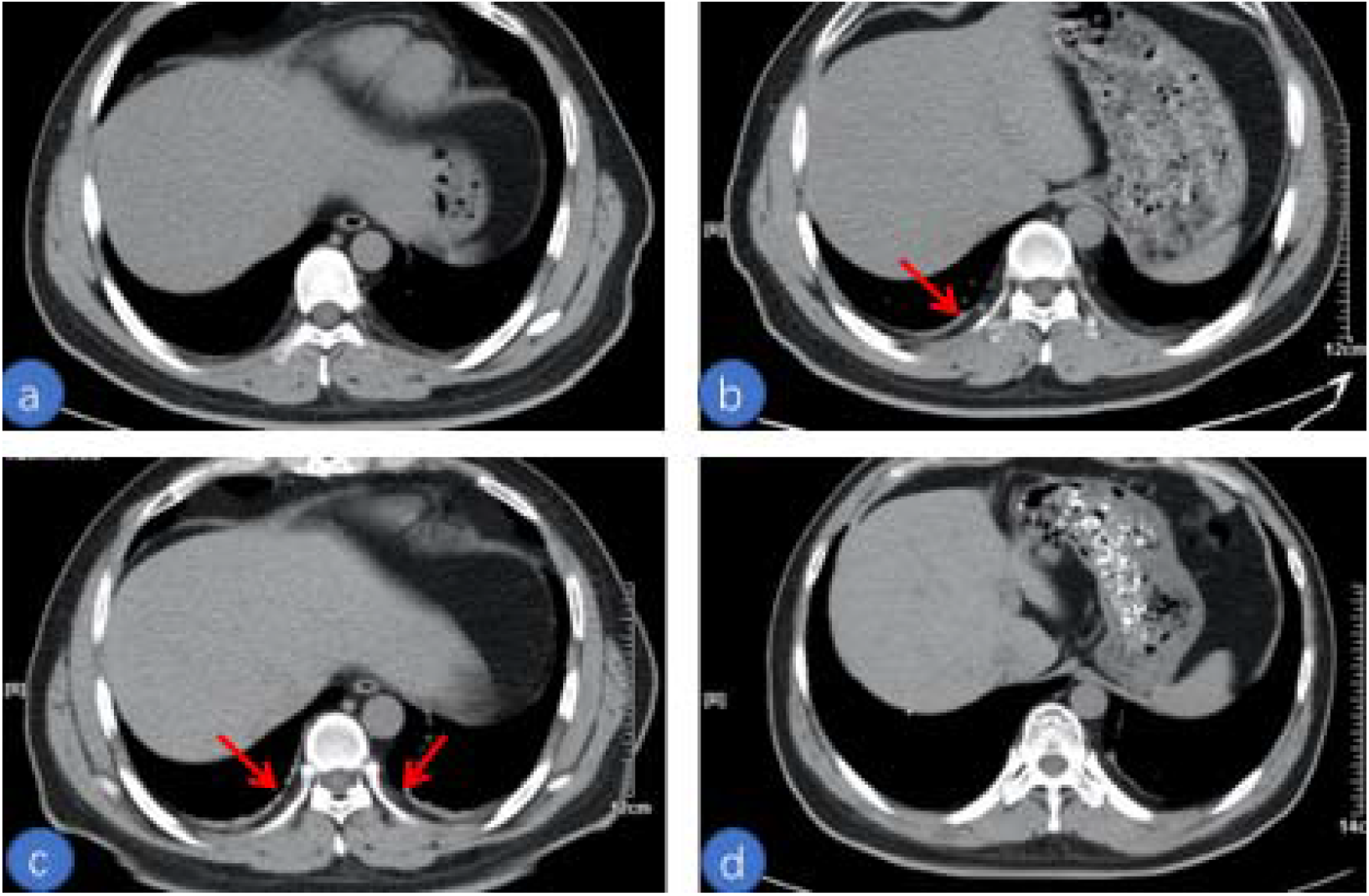
Chest CT scans (slice thickness=5mm) in a man with COVID-19 pneumonia infected while on flight. (a) Unenhanced Chest CT scan at first day after admission shows no pleural effusion in both lungs. (b) Follow-up CT scan obtained at 3 day after admission shows a small volume of pleural effusion in the pleural cavity of the right lung, but not in the left lung. (c) Follow-up CT scan obtained at 6 day after admission shows small volumes of pleural effusion in pleural cavities of both lungs. (d) Follow-up CT scan obtained at 11 day after admission shows pleural effusions in both lungs were fully absorbed.

The median number of lobes with lung opacities involvement was 4 (IQR, 1-5; range from 0 to 5). The median lung “total severity score” was 6 (IQR, 1-9; range from 0 to 13).

On admission, initial RT-PCRs on admission were positive in 9 (90%) patients, however initial chest CT were positive in only half of the ten patients. In the patient with negative initial RT-PCT, the result was positive on the third time after a week following illness onset.

### Treatment and outcomes

All patients received antiviral treatment. All received Arbidol. Lopinavir/ Ritonavir was administered to in 7 patients alongside with Darunavir/ Cobicistat in 3 patients, and alongside with Oseltamivir in 3 patients. Four patients were given empirical antibiotic treatment, 3 patients were given systematic corticosteroid treatment, and 3 patients were given oxygen support via nasal cannula. Five patients were treated with traditional Chinese Medicine (TCM). Interferon alpha inhalation and Chloroquine were not administered to our patients.

As shown in table 4, none of the patient was ever admitted to intensive care unit (ICU). Moreover, 9 (90%) patients were discharged, and one patient was still isolated in our hospital. The median hospitalized days were 20 (IQR, 13-22; ranged from 13 to 22) of the 9 discharged patients. The median MuLBSTA score was 4.5(IQR, 0-5; range from 0 to 5).

**Table 4:**
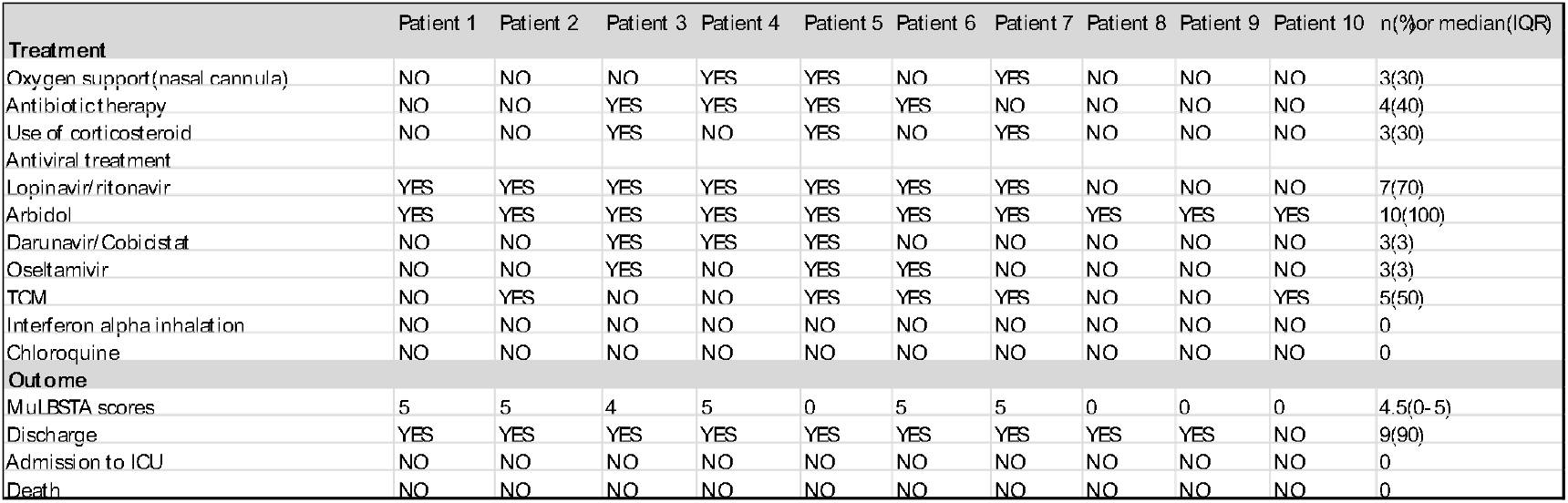
Treatment and outcomes of patients with COVID-19 by transmission on aircraft

### The risk of transmission on aircraft

To estimate the risk of aircraft transmission, we calculated the numbers of persons diagnosed with COVID-19 divided by the total number of persons on this flight, the result is 3.69% (12/325).

## Discussion

Here, we report clinical data from ten passengers with laboratory-confirmed COVID-19 after having been on a flight. As far as we know, this is the first reported case series of in-flight transmission of COVID-19. The clinical characteristics of these patients with SARS-CoV-2 transmission on aircraft were similar to those without in-flight history, as previously reported [4, 7, 14]. Notably, the symptoms of COVID-19 patients infected in this flight were relatively mild, outcomes were inclined to be better, and the risk to passengers was higher compared with transmission of SARS on aircraft [15-17].

The imaging manifestations of these patients with COVID-19 transmission on aircraft were similar to those without in-flight history, as previously reported [12]. Pleural effusion was rarely seen amongst COVID-19 patients [18, 19]. In this case, for the first time we report small volumes of pleural effusion in both lungs in a non-severe patient of COVID-19. Moreover, 9 patients were tested RT-PCR positive at admission while only 5 patients were chest CT scan positive at admission, indicating that RT-PCR has a higher sensitivity than chest CT scans in diagnosing COVID-19 in our study.

Patients in our cohort received antiviral treatment, but the types of medicine prescribed varied between patients based on their conditions. All of them received Arbidol. Use of Lopinavir combined with Ritonavia were considered to be beneficial because they had the potential to treat SARS [20]. All the rate of antibiotics, corticosteroid, and TCM use were less than half. TCM was considered to play a crucial role of treating COVID-19 patients in China, especially in the early mild period [21]. Some patients were treated with oxygen support of nasal cannula when oxygen saturations were below 93% without oxygen support and recovered. Currently, there is no widely recognized effective drug treatment exists. As, to date, no results of randomized controlled studies on these drugs in COVID-19 patients have been conducted, it was unclear whether their improvement were due to these individual drugs or some combination of them. All ten patients had no risk factors known to be associated with a worse outcome such as comorbidity or advanced age [22]. According to the currently available data, majority of COVID-19 patients infected on aircraft recovered.

We suspect the real risk of SARS-CoV-2 transmission within aircraft cabins in our investigation could be much higher than 3.69% because there might be asymptomatic COVID-19 patients among the other 313 passengers and crew members. Moreover, a period of 5 hours together in aircraft cabin with index patient is long enough to facilitate the transmission of SARS-CoV-2. Also, we believe that the risk of in-flight transmission can vary widely, influenced by whether people wore masks, physical proximity to the index patient(s), the stage of illness and numbers of index patient(s), the type of air-ventilation system, size of aircraft cabin, duration of the flight [23, 24]. The low incidences of in-flight transmission of SARS were documented in most of research reports, showing SARS-CoV was only moderately transmissible within aircraft cabins [16, 17]. However, compared with SARS, the risk of in-flight transmission for influenza appears to be far higher, with 72% of all passengers developed a clinical syndrome of influenza during the next 3 days after a 3-hour flight. Further researches are in urgent need to investigate the real magnitude and determined factors of the risk of SARS-CoV-2 transmission aboard aircraft.

We believe that a most likely time for transmission of COVID-19 in the ten passengers was during or immediately before the flight. The clustering of illness onset around 3 days in these patients is consistent with the expected incubation period of COVID-19. There was no recognized exposure history within 14 days before this fight, although it is still possible that the passengers were infected before the flight because the incubation period could be longer up to 24 days [4]. Transmission on an aircraft of SARS-CoV-2 might occur when COVID-19 patients fly, especially during the symptomatic phase of illness. Therefore, we believe that the most plausible index case resulting transmission of SARS-CoV-2 in the other nine passengers was patient 1, the 45-year-old man from Wuhan, who had onset of fever during this flight.

The COVID-19 spread rapidly around the world, largely because of infected person traveled on aircraft to other countries, since SARS-CoV-2 is more easily transmitted than SARS-CoV [25]. Different from SARS, COVID-19 can be transmitted during the incubation period [26], or by an asymptomatic patient [27]. Features of transmission between SARS and COVID-19 were largely different. For example, health workers account for majority of persons infected with SARS-CoV, while infection with SARS-CoV-2 usually develops in social clusters or family clusters [3].

Wider-Smith reported the first case in-flight transmission of SARS from Singapore [28]. They suggested that it is unlikely to have mass infection of SARS on airplanes. However, we believe it is very likely that mass infection of COVID-19 can occur during a flight, especially when respiratory and contact precautions were not in place.

How the SARS-CoV-2 in our study transmitted among the ten passengers was largely unknown. Transmission via aerosol is a possible way for SARS-CoV-2, especially when persons are placed for a long-time under high concentration of aerosol circumstances. We suspect small-particle, airborne, contact, or even large-droplet spread is the possible route of COVID-19 transmission on aircraft. To explore the potential modes of SARS-CoV-2 transmission, we might refer to anecdotal reports about other respiratory pathogens. Previous studies had documented SARS-CoV transmission in-flight was more likely developed through airborne, small-particle, close contact with index patient or contaminated fomites, while evidence of large-droplet spread was absent [15]. Instead, Mycobacterium tuberculosis and influenza virus were primarily transmitted through airborne routes [29]. Further systematic studies based on more cases infected SARS-CoV-2 within aircraft cabins are imperative to determine the exact mode of in-flight transmission.

As a lesson from SARS, three measures might decrease the risk of COVID-19 transmission on aircrafts, including substantial decrease of flights from and to the areas that are seriously affected by the COVID-19 epidemic, the implantation of strict and effective preflight screening to find out the possible symptomatic persons, and especially correct and wide use of masks by passengers and crew members during a flight [15]. Moreover, we highly appreciate and suggest the implantation of two weeks’ routine quarantine for COVID-19 after the flight when any passenger or crew member had an onset of fever or respiratory symptoms during the flight to avoid the disease being spread at the destination.

None of the nine flight attendants were symptomatic or tested to be infected although they interacted with the ill passengers during this flight, perhaps largely because of their use of masks. Therefore, we strongly suggest that mask-wearing and hand hygiene should be the imperative measures taken by passengers, flight attendants, and pilots. These measures must abide to before boarding, during the flight and after disembarkation while the global pandemic of COVID-19 is occurring.

Our study is limited by the retrospective method and sample size. Several issues should be taken into consideration when reading the results. Firstly, we tried to obtain the passenger rosters, their seats assignments, and contact information during this flight from the Scoot airline in Singapore, as well as from the Health Commission and CDC of Hangzhou, but failed. Therefore, we could not confirm that whether the transmission of COVID-19 amongst passengers occurred before boarding (having a feature that patients being randomly distributed throughout the aircraft) or while on board in the aircraft (having a feature that clustered patients were seated in the same row or within two rows in front of or behind index patient). The WHO working definition of a contact on a flight is any passenger seated in the same row of seats or within two seats in front of or behind the index patient, or any flight attendant.[30] Secondly, there were two asymptomatic patients reported in our study. We agreed that more asymptomatic carriers might be found out if all the other passengers and crew members were tested with RT-PCR assay for SARS-CoV-2 before they were released from quarantine. It was reported that presymptomatic and symptomatic patients are equally effective in spreading COVID-19.[3, 31] A possible explanation for this aircraft transmission is there was a presymptomatic or asymptomatic flight attendant who mediated transmission between passengers, although we believe it was less likely as the flight attendants were wearing masks. Thirdly, information about the physical proximity to the index patient were unavailable, therefore we could not quantify the exact risk of in-flight transmission, especially whether degree of illness in passengers was consistent with the physical proximity to the index patient(s).

SARS-CoV-2 transmission within aircraft cabins has never been addressed in all anecdotal reports on COVID-19 outbreak. It is potential for COVID-19 transmission by airplane, although the symptoms are mild. Passengers and attendants must be protected during the flight.

Systematic further investigations and more data are in urgent need to confirm the real risk, exact pattern of in-flight transmission, clinical characteristics and outcomes in COVID-19 patients infected in a flight.

## Data Availability

All data generated or analyzed during this study are included in this article.

## Disclosures of conflicts of interest

The authors of this study declare that they have no conflict of interest.

## Acknowledgements

This work was supported by Chinese Foundation for Hepatitis Prevention and Control: TianQing Liver Diseases Research Fund Subject [TQGB20150109]; Natural Science Foundation of Zhejiang Province [Q17H010001]; Natural Science Foundation of Ningbo [2017A610246]; Science and Technology Plan Project of Jiaxing City[2018AD32097]

## Notes

### Competing Interest Statement

The authors have declared no competing interest.

